# Facilitating clinical trials in Polycythemia vera (PV) by identifying patient cohorts at high near-term risk of thrombosis using rich data and machine learning

**DOI:** 10.1101/2024.01.21.24301561

**Authors:** Ghaith Abu-Zeinah, Spencer Krichevsky, Katie Erdos, Richard T. Silver, Joseph M. Scandura

## Abstract

Thrombosis remains the leading cause of morbidity and mortality for patients (pts) with polycythemia vera (PV), yet PV clinical trials are not powered to identify interventions that improve thrombosis-free survival (TFS). Such trials are infeasible in a contemporary PV cohort, even when selecting “high-risk” pts based on Age >60 and thrombosis history, because thousands of patients would be required for a short-term study to meet TFS endpoint. To address this problem, we used artificial intelligence and machine learning (ML) to dynamically predict near-term (1-year) thrombosis risk in PV pts with high sensitivity and positive predictive value (PPV) to enhance pts selection. Our automation-driven data extraction methods yielded more than 16 million data elements across 1,448 unique variables (parameters) from 11,123 clinical visits for 470 pts. Using the AutoGluon framework, the Random Forest ML classification algorithm was selected as the top performer. The full (309-parameter) model performed very well (F1=0.91, AUC=0.84) when compared with the current ELN gold-standard for thrombosis risk stratification in PV (F1=0.1, AUC=0.39). Parameter engineering, guided by Gini feature importance identified the 21 parameters (top-21) most important for accurate prediction. The top-21 parameters included known, suspected and previously unappreciated thrombosis risk factors. To identify the minimum number of parameters required for the accurate ML prediction, we tested the performance of every possible combination of 3-9 parameters from top-21 (>1.6M combinations). High-performing models (F1> 0.8) most frequently included age (continuous), time since dx, time since thrombosis, complete blood count parameters, blood type, body mass index, and JAK2 mutant allele frequency. Having trained at tested over 1.6M practical ML models with a feasible number of parameters (3-9 parameters in top-21 most predictive), it is clear that study cohorts of patients with PV at high near-term thrombosis risk can be identified with high enough sensitivity and PPV to power a clinical trial for TFS. Further validation with external, multicenter cohorts is ongoing to establish a universal ML model for PV thrombosis that would facilitate clinical trials aimed at improving TFS.

## Introduction

Polycythemia vera (PV) is chronic myeloproliferative neoplasm (MPN) characterized by increased risk for thrombosis and shortened survival. Venous thromboembolism and arterial thromboses such as myocardial infarction and strokes remain the leading causes of death in PV ^1–3^. The risk of these cardiovascular events (hereafter, collectively referred to as thrombosis) in patients with PV is at least 2-3 fold higher than individuals without PV^4^, and the age-adjusted cardiovascular mortality is almost twice that of the general population^5^. Control of excessive erythropoiesis and use of antiplatelet therapy can reduce thrombosis risk, and these interventions have been the backbone of PV therapy for many decades^1,6,7^. Nonetheless, thrombosis risk for patients with PV remains well above that observed in healthy controls, indicating that further progress must be made^3,5,8^. Yet, because the near-term thrombosis risk for most patients with PV adherent to the standard-of-care hematocrit < 45% is only ∼1-3% per year^6^, clinical trials designed to reduce thrombosis risk in PV are infeasible. Such trials would require too many patients and/or too long a follow-up. For instance, an adequately powered three-year study testing whether an intervention reduces the thrombosis rate from 2% per year to 1% per year would require almost 1,500 patients (1:1 randomization, alpha=0.05, power=0.8): a study size never accomplished in PV. Thus, inclusion criteria must be developed to select patients at greatest near-term risk for thrombosis to make such studies feasible.

Erythrocytosis, leukocytosis, inflammation, and many other factors are implicated in the increased risk of thrombosis in PV^9^. However, current risk stratification by the European Leukemia Network (ELN) and the National Comprehensive Cancer Network (NCCN) defines thrombosis risk by initial assessment of just two binary, non-modifiable risk factors (Age > 60, history of prior thrombosis) and does not usefully predict near-term thrombosis risk^10^. Given the panoply of known and unknown factors that contribute to thrombosis risk in PV, we used machine learning (ML) and high-dimensional clinical data retrieved from the electronic medical record (EMR) to identify, in an unbiased approach, the clinical parameters that best predict near-term (within 12 months) thrombosis risk. ML is a subfield of artificial intelligence (AI) that involves training algorithms to learn and identify patterns and associations within a dataset. ML algorithms are well-suited for this task because they can handle large clinical datasets with high-dimensional parameters to identify complex patterns associated with thrombosis that traditional models do not.

Rigorous parameter selection and operational testing allowed us to simplify the number, and type, of clinical parameters required for high-performing model-based predictions of near-term thrombosis risk. The path to reducing thrombosis related death and morbidity in PV will pass through clinical trials. Our ML tool allows dynamic monitoring of an individual’s thrombosis risk and is well-suited for use as an inclusion criterion that can enrich for patients at highest near-term risk and, in so doing, make feasible clinical trials to reduce thrombosis risk in a rare disease such as PV. Such a tool would be valuable in clinical practice for escalation or de-escalation of antithrombotic therapy once validated for use in clinical trials.

## Methods

### Ethics statement

The Weill Cornell Medicine Institutional Review Board (IRB) approved this study. The study was conducted in accordance with the declaration of Helsinki.

### Study design and patient population

The Richard T. Silver, M.D. Myeloproliferative Neoplasms Center (Silver MPN Center) of Weill Cornell Medicine (WCM) has developed a robust informatics infrastructure for the extraction, storage, maintenance, and aggregation of high-dimensional, longitudinal patient data from most clinical data sources. We developed MPN-centered research database repository (RDR) infrastructure that aggregates curated data from our MPN Research Electronic Data Capture (REDCap) databases, raw and research-ready data from EMRs (Epic Systems, and Allscripts Enterprise Electronic Health Records), and data from external sources (eg. CDC National Death Index) for all clinical, laboratory, and outcomes data^11^ using the Observational Medical Outcomes Partnership Common Data Model (OMOP-CDM). Patients with PV were initially identified by querying the database for International Classification of Disease (ICD) v9.0/10.0 or Systematized Nomenclature of Medicine – Clinical Terms (SNOMED-CT) codes consistent with a PV diagnosis. Diagnosis was manually validated as previously described^12^, using criteria set by the Polycythemia Vera Study Group criteria (PVSG; prior to 2007)^13^, WCM^14^, or World Health Organization (WHO) 2016 (2016-2019)^15^. Patients with non-PV diagnoses, such as secondary erythrocytosis or essential thrombocythemia, were excluded from the study. Data collection was performed until the study’s closure on April 1^st^, 2020, encompassing 470 PV patients seen at WCM as previously reported ^12^.

### Data collection and classification

Using our MPN RDR, we extracted all data available for patients with PV at each WCM Hematology-Oncology clinic visit, including all visits between initial presentation and the end of follow-up, or study closure. This “real world” dataset encompassed 11,123 visits for 470 patients^12^, and 1,448 unique parameters, yielding over 16 million data elements (Table 1).

**TABLE 1:** Demographics and clinical characteristics of retrospective PV training/testing cohort.

| Characteristic | N = 470<sup>1</sup> |
| --- | --- |
| Age | 56 (11-91) |
| Sex |  |
| Female | 229 (49%) |
| Male | 241 (51%) |
| Race |  |
| Non-White | 49 (12%) |
| White | 377 (88%) |
| Unknown | 44 |
| ELN risk |  |
| High | 302 (64%) |
| Low | 168 (36%) |
| Thrombosis history |  |
| None | 439 (93%) |
| Thrombosis | 31 (6.6%) |
| Venous thrombosis history | 17 (3.6%) |
| Arterial thrombosis history | 14 (3.0%) |
| PV duration (yrs) | 8 (0-40) |
| Follow-up at WCM (yrs) | 2 (0-17) |
| Clinic visits (n total) | 12 (1-267) |

<sup>1</sup> Median (Minimum-Maximum); n (%)

Extraction of clinical data was largely automated using our RDR and informatics workflow (Figure 1). A comprehensive list of data sources and final data elements is included in Supplementary Table 1. Structured query language (SQL) and R programming were used to extract data from the RDR including clinical data such as demographics (e.g., patient age, sex, race, ethnicity), medical history (e.g., cardiovascular disease, medications), laboratory values, bone marrow pathology findings, and gene mutations. Past medical history and adverse events (AEs) reported at each clinic visit were structured using classes and subclasses of the Common Terminology Criteria for Adverse Events (CTCAE) v5.0 ^16^ and then binarized. All non-MPN medications were extracted from the EMR medex and categorized by SNOMED (Systemized Nomenclature of Medicine – Clinical Terms) classes^17^ (Supplementary Table 1). Bone marrow findings were extracted and tabulated from clinical hematopathology reports, using a rigorously validated method of natural language processing as previously described^11^. Composite parameters, such as ELN risk (Low or High), the neutrophil-to-lymphocyte ratio (NLR)^18^, binarized age (<60 or >60 years)^19^, binarized JAK2 variant allele burden (<50% or >50%)^20^ and others were calculated from primary extracted data (e.g., laboratory values, medications, pre-existing conditions) ^18–20^ (Supplementary Table 1). Additional data was obtained from external sources through research agreements with the Centers for Disease Control (CDC) National Death Index for survival data, and a commercial laboratory for molecular data. Primary data elements such as the dates of PV diagnosis, thromboembolic events, progression to post-polycythemia vera myelofibrosis (PPVMF), or evolution to acute leukemia (AML) and the administration and duration of MPN therapies were all manually verified.

**FIGURE 1:**
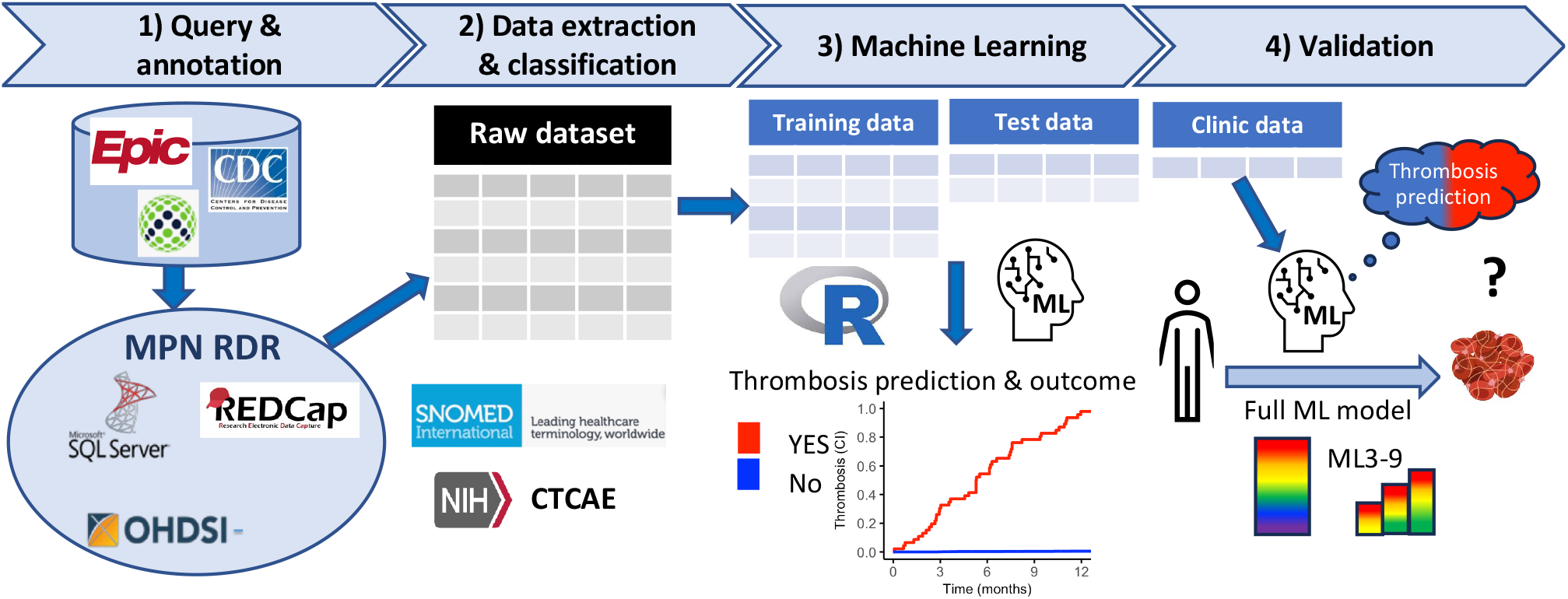
Schematic for automated data collection, retreival, and analysis pipeline.

Missing data were imputed using multivariate imputation by chained equations (MICE). We rigorously evaluated the limits of missingness that could be accommodated using MICE in the following way. We first defined a complete data set and then masked (removed) varying subsets of data up to the missingness observed within each parameter. We then imputed the masked data over 8k iterations and defined the limits of missingness tolerable by calculating the Euclidean distance and Jaccard index between partially masked/imputed and the original complete data set. Based on this analysis, we determined that parameters with <50% missingness could be imputed while maintaining a low degree of imputation error and without data corruption. Of the full list of the 309 examined parameters meeting this criterion, the majority have no missing data (n = 247, 80%, Supplementary Table 1 and Supplementary Figure 1).

### Statistical analysis and Machine Learning

Classification Machine Learning (ML) models were used to longitudinally predict, near-term thrombosis events (within 12 months) at each Hematology-Oncology clinic visit based upon the data/parameters available at the time of the visit. Our goal was to develop a predictive model that could be applied at the time of patient contact. Each clinic visit was examined separately to train dynamic models that could be applied at any time during a patient’s disease course. The AutoGluon-Tabular open-source Python package was used to evaluate several ML classification algorithms^21^. Models considered include Random Forests (RF), Extra Trees Model, eXtreme Gradient Boosting (XGBoost), Light Gradient-Boosting Machine (LightGBM), Logistic Regression (log-reg), k-nearest neighbors (kNN), and Neural Networks implemented by FastAI and Torch. These models were fine-tuned and then compared using receiving operating characteristic (ROC) analysis in an initial assessment to determine which type of model best predicted thrombosis. Based upon this analysis, RF was identified by AutoGluon as the optimal ML algorithm for predicting near-term thrombosis at each clinic visit.

Hyperparameter tuning was performed^22^ to optimize RF model training. We tuned the number of trees in the forest, the maximum number of parameters considered for splitting a node, the maximum depth (number of levels) of each decision tree, and the threshold at which prediction probabilities were discretized into positive or negative classes. We implemented a grid search technique to find the optimal combination of hyperparameters specific to each subset of parameters used for training.

Synthetic minority oversampling technique (SMOTE) was tested to reduce class imbalance since instances predictive of thrombosis were a minority. But because there was no significant difference in model performance between sampled and non-sampled data, we proceeded without oversampling to avoid model overfitting that can be introduced by SMOTE.

For RF model development, the dataset was randomly split into training (75%), and testing (25%) subsets. The testing subset was exclusively used to assess the performance of RF models trained using the training subset. F1 score, the harmonic mean of precision (positive predictive value or PPV) and recall (sensitivity) was the primary metric used to determine model performance but additional performance metrics—accuracy, sensitivity, specificity, PPV, negative predictive value (NPV), and area under the curve (AUC)—were also evaluated.

Because requiring all 309 variables in a ML model (“ML Full Model”) is not practical for clinical use, we rigorously sought to identify the highest-performing models requiring the fewest parameters. We first identified the top 21 parameters (top-21) most predictive of near-term thrombosis in the ML Full Model using Gini feature importance^23^. To further reduce the number of parameters, we tested the performance of models derived from every combination of the top-21 in groups of 3 through 9 parameters. A total of >1.6M models were trained and tested for performance (see F1 bean plot Figure 4A). To optimally enrich for patients with high near-term thrombosis risk, we identified models with high F1 score > 0.8 and an optimal balance of sensitivity and PPV based on a feasibility analysis for complete clinical trial accrual. A trial was considered feasible if “time to accrual” was predicted to be less than 3 years. In this analysis, “time to accrue” was estimated from both the sample size required to meet 50% reduction of thrombosis in a 1:1 randomized clinical trial with α 0.05 and power of 0.8 (calculated based on model’s PPV) and the time required for eligible patients to be identified (calculated based on sensitivity and the number of PV patients seen in WCM clinic).

Statistical analyses were performed in RStudio software and utilized associated packages (Supplementary References). Descriptive and summary statistics using Analysis of Variance (ANOVA), Student t-tests, Mann-Whitney tests, and Fisher’s tests were employed to compare the difference between groups. The cumulative incidence (CI) of thrombotic events was generated by Kaplan-Meier analyses compared between risk groups using Fine-Gray models. The timepoint corresponding to the greatest change in CI trajectory was identified via bilinear fitting using custom scripts.

## Results

Of the 470 patients in the PV cohort, 409 had longitudinal follow-up of at least 12 months during the study period for inclusion in our ML analysis. The median age of the 470 patient cohort at diagnosis was 56 (range 11-91) and 229 (49%) were females (Table 1). Median duration from diagnosis was 8 years (range 0-40) and median follow-up at WCM was 2.2 years (range 0-17). History of thrombosis was present at diagnosis in 31 (6.6%), and 302 (64%) were high-risk by ELN/NCCN criteria (Table 1). Over the course of WCM follow-up, 72 thrombotic events (39 venous, 33 arterial) occurred in 60 patients (Supplementary Figure 2), an incidence rate (IR) of 2.55 events/100 patient-years. There was no significant difference between the cumulative incidence (CI) of arterial and venous events (Supplementary Figure 3). The CI of thrombosis after diagnosis was non-linear (Figure 2A), with the rate of thrombosis much higher within the first ∼2 years of diagnosis compared to subsequent years (IR 4.4% vs 1%, respectively). Similarly, the CI of recurrent thrombosis after an initial event was also much higher in the first ∼2 years after the thrombotic event (IR 9.3% vs 1.3%, Figure 2B).

**FIGURE 2:**
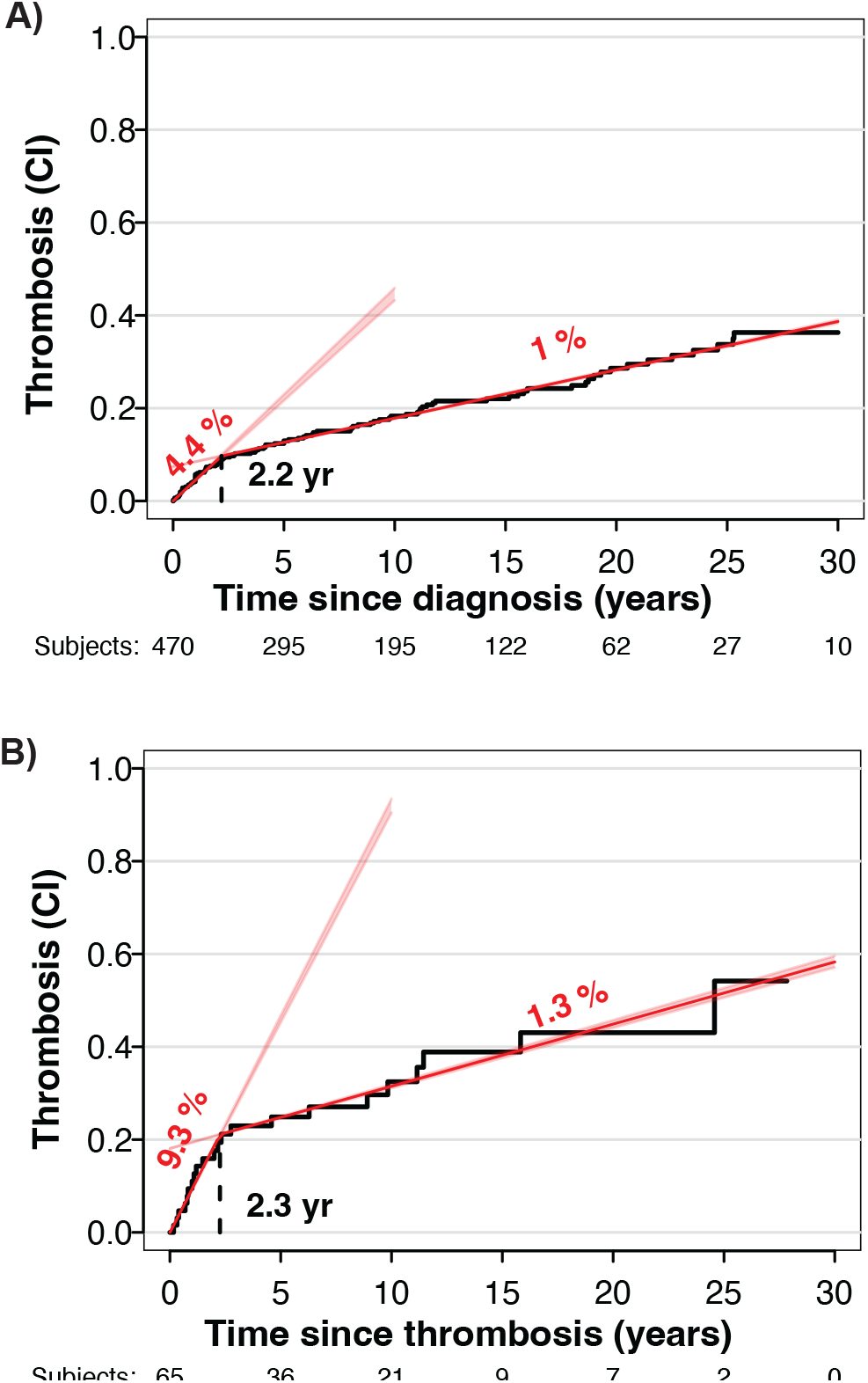
Cumulative incidence of thrombosis from the time of A) Diagnosis and B) first thrombosis.

We trained a RF ML model to predict near-term thrombosis risk (within 12 months) using all 309 parameters available at each clinic visit for the 306 patients in the training subset (Figure 3A). Predictions were made at each clinic visit based upon data available at that time. This “RF Full Model” was then applied to data from the 103 patients in the test subset to evaluate performance. The RF Full Model was highly predictive of thrombosis with F1 of 0.89 (Accuracy=99%, sensitivity=86%, specificity=100%, PPV=83%, NPV=100%). Most clinic visits classified as high risk by RF Full Model (RF-high) were followed by a thrombosis event within 12 months (Figure 3B), In contrast, only 9 out of 2044 (0.44%) clinic visits classified as low risk by RF Full Model (RF-low) were followed by a thrombosis, a statistic comparable to the annual incidence of venous and arterial thromboses combined, in the general population^24,25^. In ROC curve analysis, the AUC of the ML Full Model indicated excellent performance whereas ELN Risk performed poorly as a near-term predictor of thrombosis (0.92 versus 0.64, p<0.01) (Figure 3C).

**FIGURE 3:**
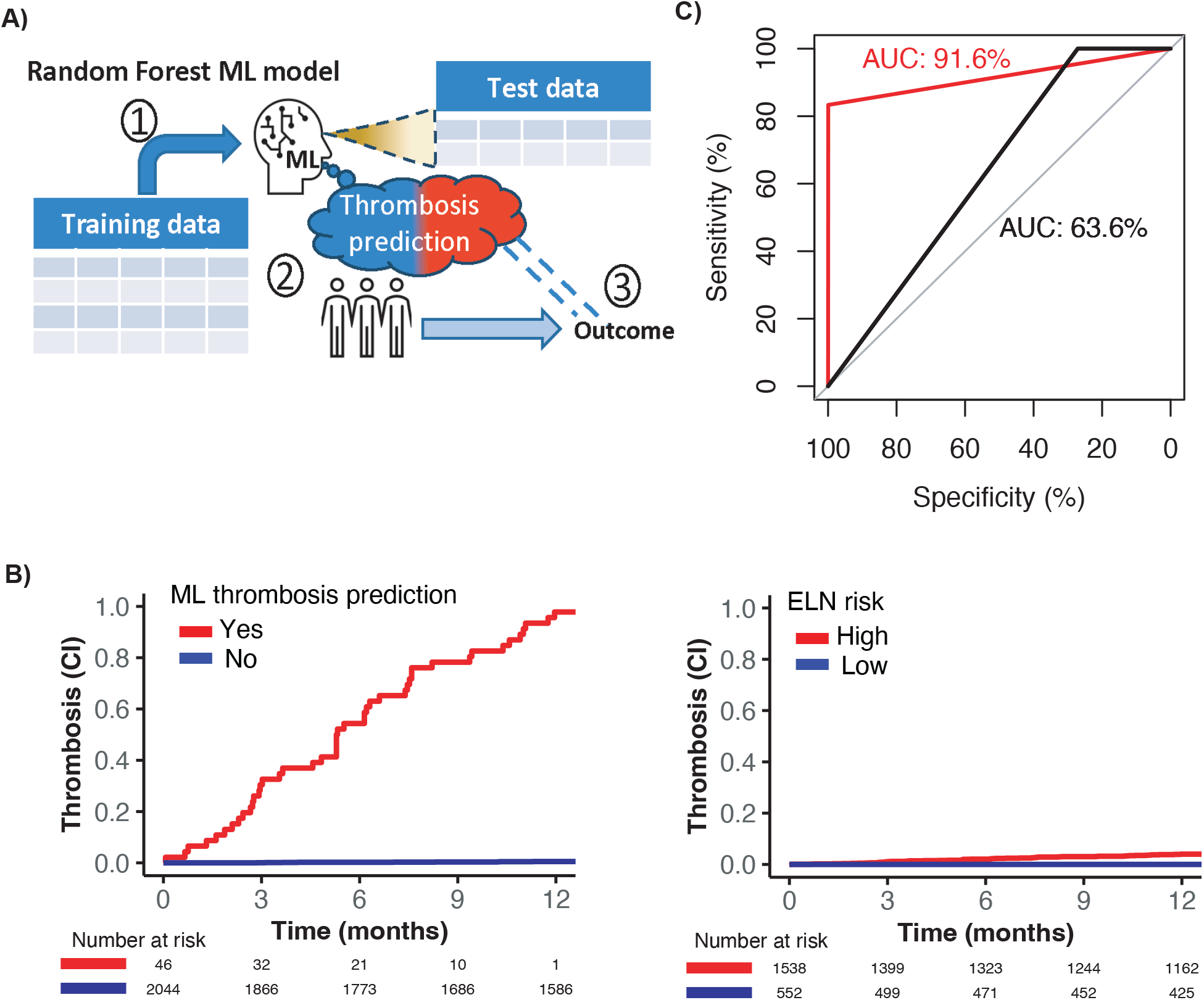
Machine learning (ML) Random Forests (RF) approach. predicts patient risk on each clinic visit nlikely to have thrombosis based on clinical data (A), and provides accurate prediction of throm-nce within a year for patients in the test subset (B), outperforming ELN risk prediction with greater

We identified the top 21 parameters (top-21) most predictive of near-term thrombosis risk in the ML Full Model using Gini feature importance (Supplementary Table 2 and Supplementary Figure 4). Several parameters within this list have been previously implicated in PV thrombosis (e.g., age, blood cell counts, JAK2 allele burden). Time since diagnosis (Time_since_dx) and time since last thrombosis (Time_since_thromb) were also identified within the top-21 parameters, as expected from the non-linear cumulative incidence of thrombosis plots (Figure 2) and prior reports^8^. The top-21 parameters also included known thrombosis risk factors that have not been previously linked to PV thrombosis risk (e.g., blood type^26,27^, BMI, creatinine). Finally, the top-21 list included some parameters with previously unrecognized contributions to thrombosis risk (MCV, uric acid, LDH) and, for that reason, not previously assessed in traditional analyses. Most, but not all, of the top-21 parameters had a linear association with thrombosis events (Supplementary Table 3).

Clinical application of the RF Full Model would not be possible without the robust research infrastructure available through our RDR. To overcome this barrier for other centers, we worked to identify high performing models that required a minimal number of readily available clinical variables. We focused on parameters within the top-21 and evaluated the performance of >1.6 million ML models comprising every combination of 3 to 9 parameters (ML3-9) (Figure 4A). Several trends emerged from this analysis. Firstly, models with more parameters had a higher median performance, but the main driver was a reduction in the proportion of models with poor performance. Nonetheless, even the best models comprised of just 3 or 4 parameters underperformed in comparison to models that considered more parameters. Second, the proportion of models with excellent performance (F1> 0.8) was not significantly better for models comprised of 7, 8 or 9 parameters compared to those assessing only 5, suggesting that the additional complexity of the higher parameter models did not guarantee better performance. We concluded that top performing RF ML models with 5-7 parameters provide the best balance between performance and practicality.

**FIGURE 4:**
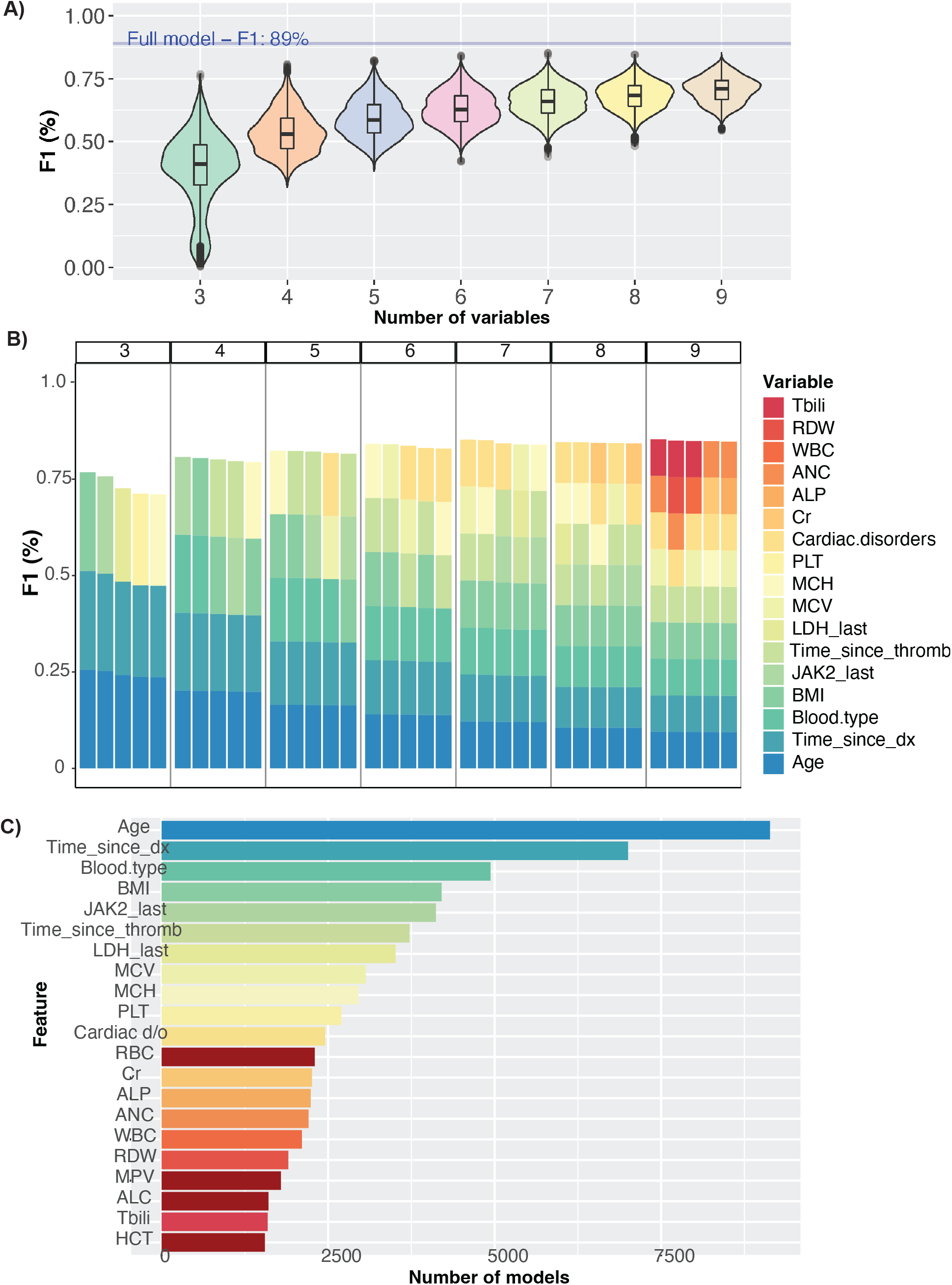
dentification of best performing ML models with 3-9 variables (ML3-9) from the Gini feature top 20. A) F1 for ML3-9 with reference to full ML F1. B) Samples of top performing ML3-9. C) Variables most frequently included in top performing ML3-9

We discovered that some parameters were repeatedly present in the best performing models. For instance, the best 3 parameter models always included Age (continuous) and time since diagnosis (Time_since_dx) alongside a varying third parameter (Figure 4B). We analyzed these trends systematically, determining the frequency by which each parameter was found among the best performing models (Figure 4C). Age (continuous), Time_since_dx, blood type, body mass index (BMI), JAK2 allele frequency, time since thrombosis (Time_since_thromb), and LDH were the parameters most represented in high performing models. All the other top20 parameters also appeared in some models and combinations suggesting that many potential models could be used to successfully predict near-term thrombosis.

A model with F1> 0.8 alone does not necessarily guarantee that patient selection for a thrombosis-prevention clinical trial will be feasible. For instance, if too selective, it may take a very long time to identify patients, but if not selective enough, many patients will be required for statistical power. Therefore, we performed a feasibility analysis of time to enrollment using model sensitivity and PPV – performance metrics that determine F1. Higher sensitivity models are more inclusive or can identify more of the patients at high near-term risk. This translates to shorter time required to find an eligible patient for a clinical trial (Supplementary Figure 5A). But high sensitivity needs to be coupled with higher PPV that guarantees a smaller sample size required for a clinical trial to meet its endpoint (Supplementary Figure 5A-B). A balance of higher sensitivity and sufficient PPV together are associated with shorter time to study enrollment and completion (Figure 5).

**FIGURE 5:**
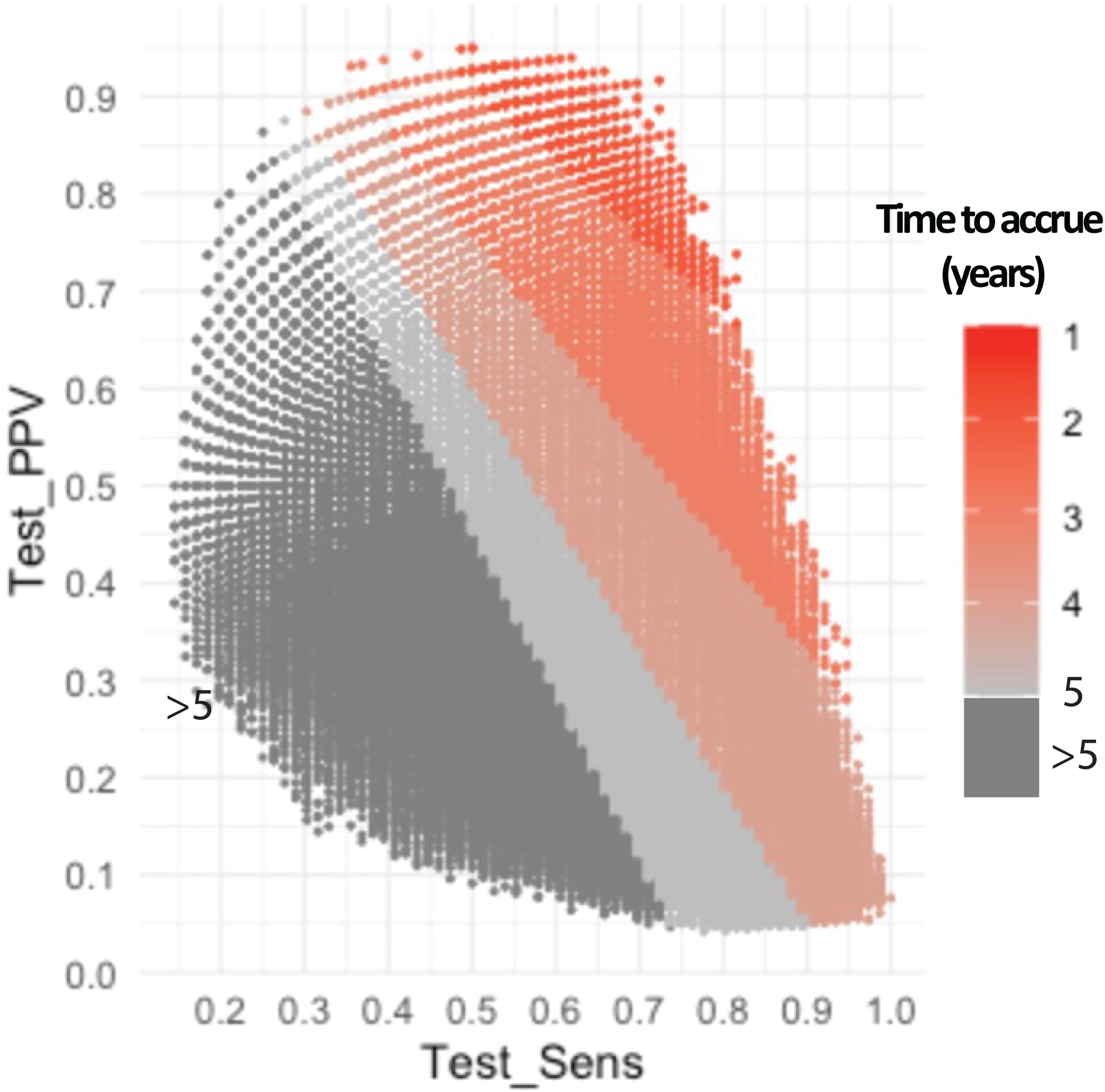
Years to accrue PV patients to a clinical trial of throm-bosis prevention according to ML3-9 model sensitivity and PPV.

## Discussion

Thrombosis remains a leading cause of morbidity and death associated with PV and no methods exist to identify the patients at highest acute risk. Part of the challenge in risk assessment is that individual risk is not uniform across all patients and is not linear over time. Currently, two binary, non-modifiable factors (age and thrombosis history) are used by ELN/NCCN to consider patients with PV either as “low risk” or “high risk” for thrombosis. Although over time, the “high risk” group experiences more thrombotic events than the “low risk” group, both experience far more events than expected in healthy controls thus indicating that “low risk” is a misnomer. Conversely, most patients with “high risk” PV never experience a thrombotic event after starting treatment indicating that the “high risk” is acutely high in only a subset of these patients. Part of this deficiency likely stems from the invariant, linear rate of thrombosis assumed by the ELN/NCCN model that is not observed clinically (Figure 2)^8^. This study accommodates the observed non-linear incidence of thrombosis, allowing dynamic prediction that accounts for changing risk, which linear models fail to do. A major limitation of the ELN/NCCN criteria is that it performs poorly at predicting near-term thrombosis (Figure 3), therefore limiting its usefulness as a clinical trial inclusion criterion.

This work used machine learning (ML) to identify individualized thrombosis risk for patients with PV. ML models thrive in data-rich environments. We developed tools to extract and analyze high-dimensional data available in the EMR and allow unbiased identification of clinical factors most predictive for thrombosis. Critically, we included all available data rather than selecting variables based on pre-supposed importance as a means for discovering previously unconsidered variables. This proved to be critical as several important parameters we identified have not be previously implicated in PV thrombosis risk. Serial data was important because the risk of thrombosis varies significantly with time^8^ (Figure 2). Within the first two years of diagnosis, or following a clot, the rate of thrombosis can be almost 10-times higher than the overall thrombosis rate. Thus, training the ML model using serial data was necessary to accommodate the obvious non-linearity of thrombosis risk.

Over 300 clinical parameters were assessed to identify those most helpful in predicting thrombosis within 12 months. Among the most important markers were several factors that have been previously implicated in prognosis of PV thrombosis such as age (continuous, but not binary <60/ ≥60)^19^, blood counts, history of prior clots and JAK2 allele burden (continuous, but not binary <50%/ ≥50%)^20^. Importantly, we found that age was more predictive when treated as a continuous variable rather than as a binary discriminator. The temporal nature of risk was also evident where time since most recent thrombosis was more important than the binary history of prior thrombosis. This finding is evident in the cumulative incidence plots because the recurrent thrombosis rate is very high only during the first two years following a thrombotic event but subsequently decreases to a rate of just over 1% per year (Figure 2). Clinicians may recognize that thrombosis risk is higher around the time of PV diagnosis and the ML models identified time since diagnosis among the most important parameters predicting thrombosis risk. It is noteworthy that most of these important parameters predicting thrombosis in PV are either defined by time or known to vary with time.

The broad, unbiased ML analysis performed also identified additional thrombosis predictors that have not been previously considered in PV. ML identified non-O blood type as a risk factor for thrombosis in patients with PV (Figure 4). Non-O blood type was associated with a 1.5-fold higher risk for thrombosis compared to O blood type (p=0.001, Supplementary Table 3). These results align well with large population studies that have shown that ABO blood type is highly associated with cardiovascular events both in healthy controls and in patients with cancer^26,27^. ML also established thrombosis risks due to BMI and creatinine; both factors that have been implicated in overall thrombosis risk but not previously considered in PV thrombosis risk assessment^28,29^. ML also identified other risks for thrombosis in PV (e.g., LDH, MCV, and uric acid) that have not been previously identified as thrombosis risk factors in PV or in other clinical settings. These data suggest that PV thrombosis risk is a composite of both permanent and dynamic clinical parameters.

The ML Full Model encompassed > 300 variables was highly predictive for thrombosis within 1 year, but such a data-rich model would have been infeasible without a robust health informatic infrastructure. To identify simplified models that can be more easily applied in clinical practice, we trained and tested over 1.6 million models containing every combination of 3 to 9 of the most important clinical parameters (top-21) identified by the full model (Figure 4). This analysis demonstrated that Age, time since diagnosis, Blood Type, BMI, most recent *JAK2*^*V617F*^ allele frequency, and time since thrombosis repeatedly appear in the best performing simplified models. A model comprised of these six variables performed at a high level (F1=0.8, Sensitivity = 0.8, Specificity =0.99, PPV = 0.8, NPV=0.99, and Accuracy=0.99). Yet, other simplified models comprised of some but not all these six parameters admixed with other parameters also have a high performance. This indicates that classifying PV thrombosis using ML allows some flexibility as to the type of data required for accurate near-term clot prediction.

One of our goals was to identify patients at highest near-term thrombosis risk so that randomized clinical trials (RCT) could be performed to test new approaches to improve thrombosis-free survival (TFS) in PV. To do this, we assessed the feasibility of running such a study by estimating the time it would take to accrue to a RCT (1:1 randomization) powered to identify a 50% reduction in thrombosis risk due to a study intervention vs standard of care in patients who are otherwise well managed using anti-platelet agents and interventions to control erythrocytosis. In doing so, we realized that we need to balance the time required to accrue to a study (related to frequency with which these patients exist in our cohort) against the time it would take to observe enough thromboses to power the study (related to thrombosis rate in the RF-high group, which is essentially a model’s PPV). Feasibility analysis (see methods) was performed to identify “high-performing” models with the shortest time to accrue patients. Using the example of a high-performing six-parameter model (Age, Time_since_dx, Blood Type, BMI, most recent *JAK2*^*V617F*^allele frequency and Time_since_thromb), we calculate that at our center a fully powered study would require 45 RF-high patients and could be accrued within a ∼3 year time period. In contrast, using ELN “high risk” PV as an inclusion criterion for our hypothetical 3-year study would require over 3000 patients to establish a 50% risk reduction because these patients, considering optimal HCT control < 45%, have a yearly thrombosis risk of approximately 1% per year^6^. Such as study using ELN/NCCN criteria is entirely infeasible.

This study’s novel findings suggest that PV thrombosis risk is non-linear and represents a composite of many more factors than age and thrombosis history. The strengths of this study begin with the informatic infrastructure that facilitates extraction of all available clinical data for training. The approach of providing the model with all available data is unique because it minimizes bias that is typically introduced by selection of data variables for which to train the model. The most important variables are ultimately selected based on their availability and contribution to risk prediction and not based on the researcher or clinician’s preference. Additional strengths include utilization of AI for the comprehensive assessment of over 1.6M models for unbiased model selection. A weakness we acknowledge is the training of the data using a single center cohort at a specialty MPN center, which may not be representative of PV patients in the general community. Further validation with external and multicenter cohorts is ongoing to establish a universal ML model for PV thrombosis risk as a patient selection tool for clinical trials aimed at improving TFS/OS and in potentially in clinical practice.

## Data Availability

All data produced in the present study are available upon reasonable request to the authors

## Acknowledgements

We thank members and the leadership of the Weill Cornell Medicine ARCH team for a productive collaboration in developing the informatic tools and RDR that enabled this study.

## Notes

### Competing Interest Statement

The authors have declared no competing interest.

### Funding Statement

This study did not receive any funding

### Author Declarations

The Institutional Review Board of Weill Cornell Medicine approved this study. Protocol # 19-12021151

## References

1. Dameshek W. Physiopathology and course of polycythemia vera as related to therapy. J. Am. Med. Assoc. 1950;142(11):790–797.

2. Chievitz E, Thiede T. Complications and Causes of Death in Polycythaemia Vera. Acta Med. Scand. 1962;172(5):513–523.

3. Stein BL, Patel K, Scherber RM, et al. Mortality and Causes of Death of Patients with Polycythemia Vera: Analysis of the Reveal Prospective, Observational Study. Blood. 2020;136(Supplement 1):36–37.

4. Barbui T, Masciulli A, Carobbio A, et al. Ropeginterferon alfa-2b versus phlebotomy in low-risk patients with polycythaemia vera (Low-PV study): a multicentre, randomised phase 2 trial. Artic. Lancet Haematol. 2021;8:175–84.

5. Abu-Zeinah K, Saadeh K, Silver RT, Scandura JM, Abu-Zeinah G. Excess mortality in younger patients with myeloproliferative neoplasms. Leuk. Lymphoma. 2022;1–5.

6. Marchioli R, Finazzi G, Specchia G, et al. Cardiovascular Events and Intensity of Treatment in Polycythemia Vera. N. Engl. J. Med. 2013;368(1):22–33.

7. Landolfi R, Marchioli R, Kutti J, et al. Efficacy and Safety of Low-Dose Aspirin in Polycythemia Vera. N. Engl. J. Med. 2004;350(2):114–124.

8. Hultcrantz M, Björkholm M, Dickman PW, et al. Risk for Arterial and Venous Thrombosis in Patients With Myeloproliferative Neoplasms: A Population-Based Cohort Study. Ann. Intern. Med. 2018;168(5):317–325.

9. Hasselbalch HC, Elvers M, Schafer AI. The pathobiology of thrombosis, microvascular disease, and hemorrhage in the myeloproliferative neoplasms. Blood. 2021;137(16):2152–2160.

10. Barbui T, Tefferi A, Vannucchi AM, et al. Philadelphia chromosome-negative classical myeloproliferative neoplasms: revised management recommendations from European LeukemiaNet. Leukemia. 2018;32(5):1057–1069.

11. Sholle E, Krichevsky S, Scandura J, Sosner C, Campion T. Lessons Learned in the Development of a Computable Phenotype for Response in Myeloproliferative Neoplasms. IEEE Int. Conf. Healthc. Informatics. IEEE Int. Conf. Healthc. Informatics. 2018;2018:328.

12. Abu-Zeinah G, Krichevsky S, Cruz T, et al. Interferon-alpha for treating polycythemia vera yields improved myelofibrosis-free and overall survival. Leuk. 2021 359. 2021;35(9):2592–2601.

13. Berlin NI. Diagnosis and classification of the polycythemias. Semin. Hematol. 1975;12(4):339–51.

14. Silver RT, Chow W, Orazi A, Arles SP, Goldsmith SJ. Evaluation of WHO criteria for diagnosis of polycythemia vera: a prospective analysis. Blood. 2013;122(11):1881–6.

15. Arber DA, Orazi A, Hasserjian R, et al. The 2016 revision to the World Health Organization classification of myeloid neoplasms and acute leukemia. Blood. 2016;127(20):2391–405.

16. Cancer Institute N. Common Terminology Criteria for Adverse Events (CTCAE) Common Terminology Criteria for Adverse Events (CTCAE) v5.0. 2017.

17. SNOMED Clinical Terms. U.S. Natl. Libr. Med. 2022;

18. Carobbio A, Vannucchi AM, De Stefano V, et al. Neutrophil-to-lymphocyte ratio is a novel predictor of venous thrombosis in polycythemia vera. Blood Cancer J. 2022 122. 2022;12(2):1–6.

19. Marchioli R, Finazzi G, Landolfi R, et al. Vascular and neoplastic risk in a large cohort of patients with polycythemia vera. J. Clin. Oncol. 2005;23(10):2224–2232.

20. Guglielmelli P, Loscocco GG, Mannarelli C, et al. JAK2V617F variant allele frequency >50% identifies patients with polycythemia vera at high risk for venous thrombosis. Blood Cancer J. 2021;11:.

21. Erickson N, Mueller J, Shirkov A, et al. AutoGluon-Tabular: Robust and Accurate AutoML for Structured Data. 2020;

22. Probst P, Wright M, Boulesteix A-L. Hyperparameters and Tuning Strategies for Random Forest. 2019;

23. Menze BH, Kelm BM, Masuch R, et al. A comparison of random forest and its Gini importance with standard chemometric methods for the feature selection and classification of spectral data. BMC Bioinformatics. 2009;10(1):1–16.

24. Næss IA, Christiansen SC, Romundstad P, et al. Incidence and mortality of venous thrombosis: a population-based study. J. Thromb. Haemost. 2007;5(4):692–699.

25. Yeh RW, Sidney S, Chandra M, et al. Population Trends in the Incidence and Outcomes of Acute Myocardial Infarction. N. Engl. J. Med. 2010;362(23):2155–2165.

26. Groot HE, Sierra LEV, Said MA, et al. Genetically determined ABO blood group and its associations with health and disease. Arterioscler. Thromb. Vasc. Biol. 2020;40:830–838.

27. Englisch C, Moik F, Nopp S, et al. ABO blood group type and risk of venous thromboembolism in patients with cancer. Blood Adv. 2022;6(24):6274–6281.

28. Gecht J, Tsoukakis I, Kricheldorf K, et al. Kidney dysfunction is associated with thrombosis and disease severity in myeloproliferative neoplasms: Implications from the german study group for mpn bioregistry. Cancers (Basel). 2021;13(16):4086.

29. Samad F, Ruf W. Inflammation, obesity, and thrombosis. Blood. 2013;122(20):3415–3422.

