## Supplementary Figures for "Facilitating clinical trials in Polycythemia vera (PV) by identifying patient cohorts at high near-term risk of thrombosis using rich data and machine learning"

**<u>SUPPLEMENTARY FIGURE 1:</u> Missing data across variables**

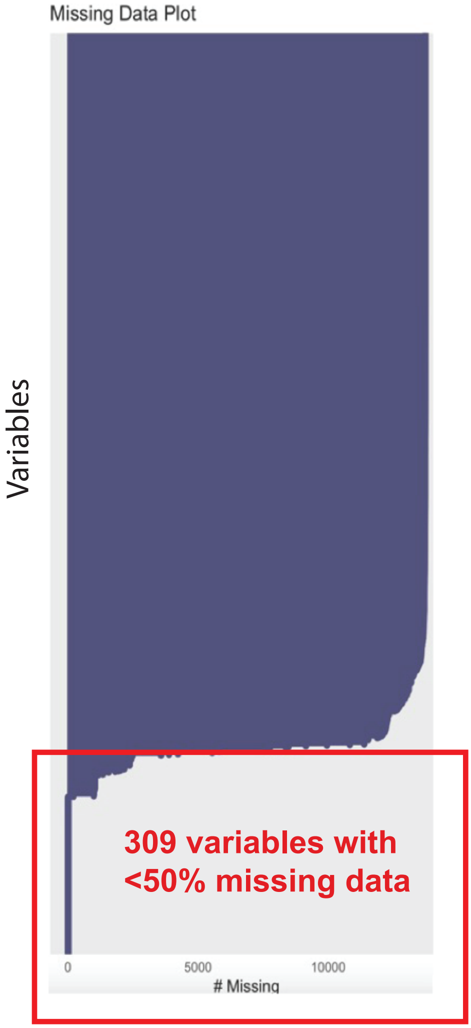


**<u>SUPPLEMENTARY FIGURE 1:</u> Swimmer plot showing thrombosis events in PV patients during WCM follow-up (red circles, blue intervals) and prior to WCM visits (yellow circles, gray intervals).**

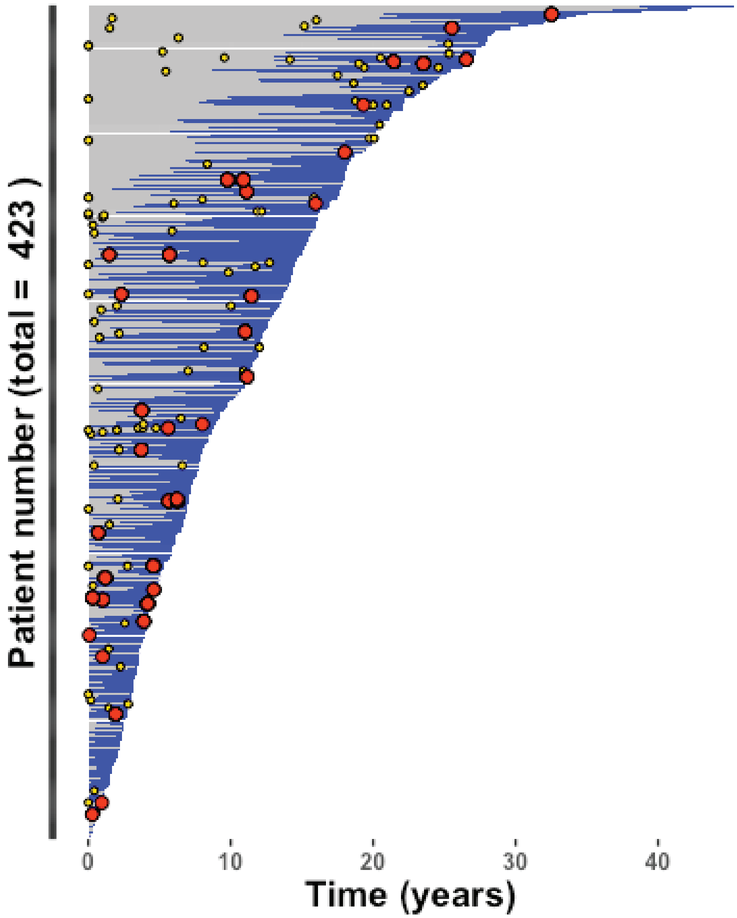

**SUPPLEMENTARY FIGURE 2:** **Cumulative incidence of thrombosis (black), stratified by venous (blue) and arterial (red) from the time of A) Diagnosis and B) first thrombosis**

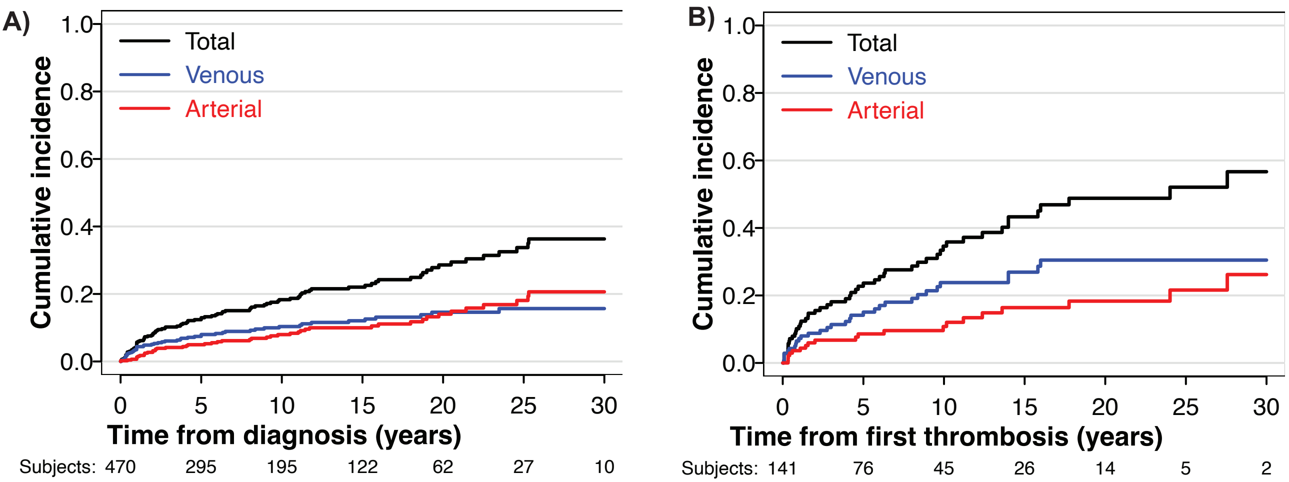


**SUPPLEMENTARY FIGURE 3:** Gini feature importance of RF ML ranking top variables.

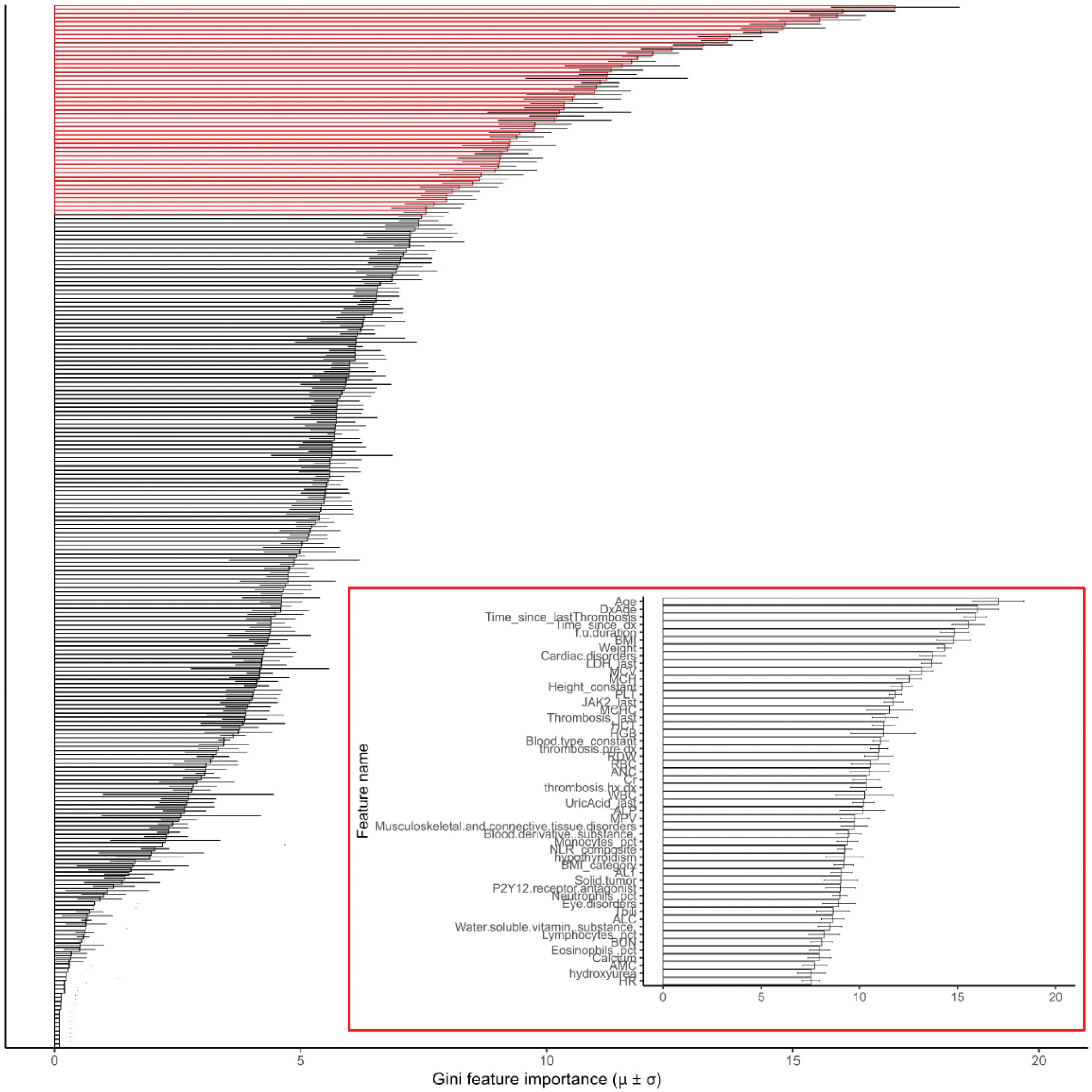

**SUPPLEMENTARY FIGURE 4:** Sample size and time to accrue to a clinical trial in PV preventing thrombosis is dependent on selection model's positivie predictive value (PPV) and sensitivity as shown in the schematic (A) of hypothetical scenarios with varying sensitivity, PPV and in (B) showing estimated sample size required according to prediction model's PPV.

A)

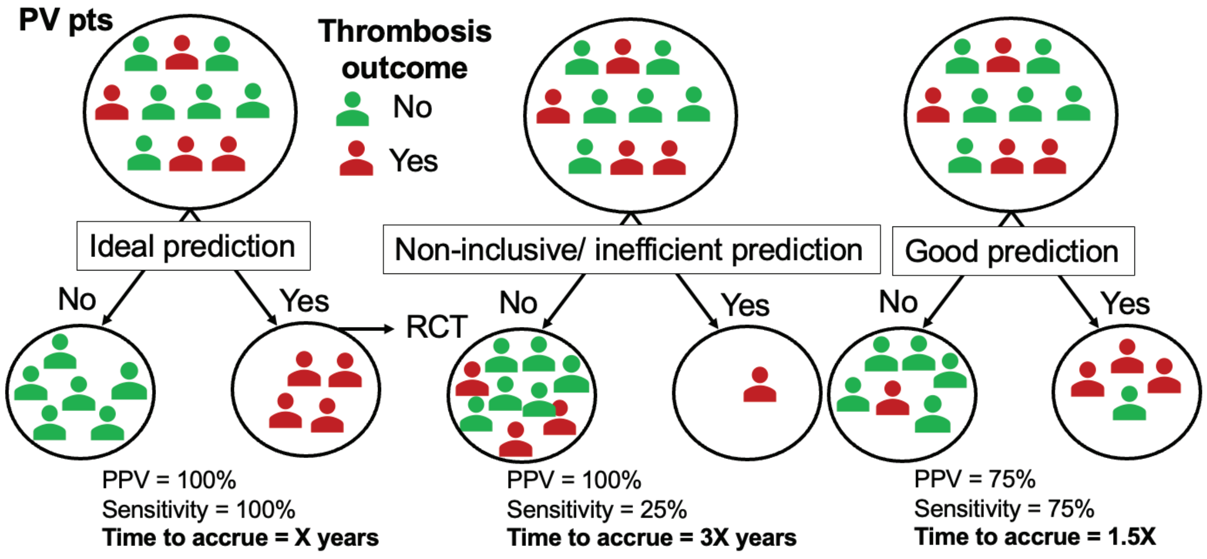


B)

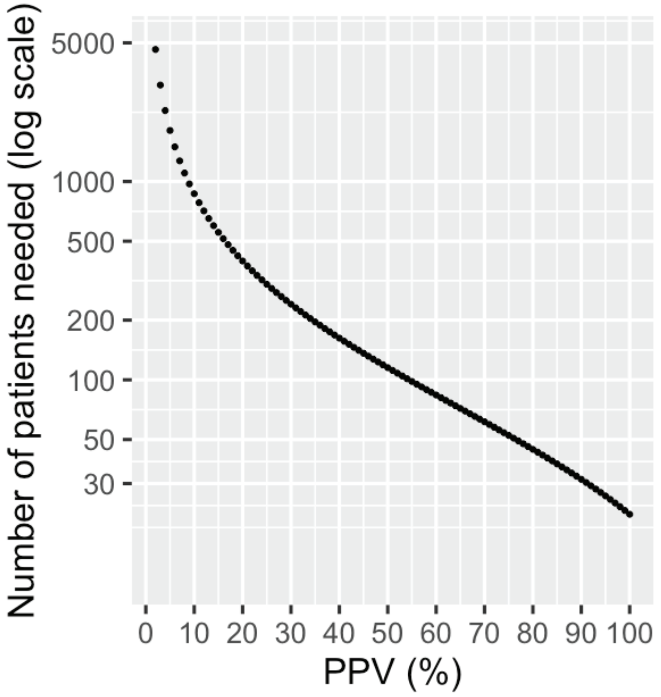
